# Randomized Clinical Trials or Convenient Controls: TREWS or FALSE?

**DOI:** 10.1101/2022.08.08.22278526

**Authors:** Shamim Nemati, Supreeth P. Shashikumar, Andre L. Holder, Gabriel Wardi, Robert L. Owens

## Abstract

We read with interest the Adams et al.^1^ report of the TREWS machine learning (ML)-based sepsis early warning system. The authors conclude that large-scale randomized trials are needed to confirm their observations, but assert that their findings indicate the potential for the TREWS system to identify sepsis patients early and improve patient outcomes, including a significant decrease in mortality. However, this conclusion is based upon a comparison of those whose alert was confirmed vs. not confirmed within 3 hours, rather than random allocation to TREWS vs. no TREWS. Using data from over 650,000 patient encounters across two distinct healthcare systems, we show that the findings of Adams et al. are likely to be severely biased due to the failure to adjust for ‘processes of care’-related confounding factors.

---

While Adams et al. do not explicitly state the mechanism by which the study intervention improves outcomes, they presumably believe one of the following to be responsible: 1) early recognition of sepsis provided by the TREWS alert followed by clinician acknowledgement and patient evaluation, 2) a streamlined user interface (UI) for guiding caregivers through a protocolized sepsis care bundle^4^, or 3) the combined effect of early recognition and patient evaluation, referred to as human–machine teaming^3^. We have reproduced their cohort characteristics in Figure 1. As can be seen, the data suggesting value to the implementation of the TREWS algorithm comes from a comparison of groups A (evaluated with 3 hours of alert) and B (evaluated more than 3 hours after the alert). Using our representation of the data, the number of false alarms (D) and missed detections (E and potentially C) becomes apparent. One even might argue for the group of septic patients not flagged by the alert (group E and potentially C) as the control group. Notably, both these groups had lower unadjusted mortality rates compared to the study arm (See Adams et al., Table 1). Nevertheless, we aimed to determine whether other factors might explain the apparent success of the algorithm within groups A relative to group B.

**Table 1.** Cohort statistics

|  | **Cohort A**<br>231,441 ED encounters (8351 septic) |  |  | **Cohort B**<br>431,780 ED encounters (23,120 septic) |  |  |
| --- | --- | --- | --- | --- | --- | --- |
|  | **Case**<br>(658) | **Control**<br>(2745) | **P value** | **Case**<br>(1614) | **Control**<br>(5942) | **P value** |
| Age (years) | 53.5 (±16.7) | 55.0 (±17.2) | 0.1 | 57.1 (±17.8) | 58.8 (±17.3) | < 0.01 |
| Documented men, N (%) | 376 (57.1%) | 1587 (57.8%) | 0.75 | 877 (54%) | 3321 (56%) | 0.2 |
| Mean CCI | 1.9 (±1.6) | 2.1 (±1.6) | 0.02 | 5.7 (±3.8) | 5.9 (±3.7) | <0.05 |
| SOFA | 1.3 (±1.9) | 1.4 (±1.9) | 0.6 | 1.0 (±1.6) | 1.3 (±1.7) | < 0.01 |
| Serum lactate | 3.0 (±1.5) | 3.0 (±1.7) | 0.6 | 2.5 (±1.3) | 2.3 (±1.4) | <0.01 |
| Serum lactate > 2 mmol/L (%)<sup>†</sup> | 82.2% | 11.9% | – | 64.9% | 11.9% | – |
| Time to antibiotics (h) | **2.2**<br>**[0.9 5.5]** | **4.6**<br>**[2.2 9.0]** | <0.001 | **1.7**<br>**[0.7 4.5]** | **3.3**<br>**[1.6 6.5]** | <0.001 |
| In-hospital mortality, n (%) | **22 (3.3%)** | **176 (6.4%)** | <0.001* | **58 (3.6%)** | **303 (5.1%)** | <0.005* |
| Maximum SOFA over 72 h | 2.7 | 3.8 | < 0.001 | 1.6 | 2.4 | < 0.001 |
| Length of stay (h) | 96<br>[48 240] | 144<br>[72 294] | < 0.001 | 118<br>[69 221] | 123<br>[68 230] | 0.06 |

<sup>†</sup> The denominator is the total number of patients in each group and therefore affected by the number of missing lactates.
\* This is the p value after adjustment for Age, Gender, CCI, Lactate, SIRS, SOFA, and an indicator for belonging to the control arm. The adjusted odds ratios for in-hospital mortality for the control arms in cohorts A and B were 2.21 and 1.6. SOFA = sequential organ failure assessment, CCI = Charlson Comorbidity Index.

**Figure 1.**
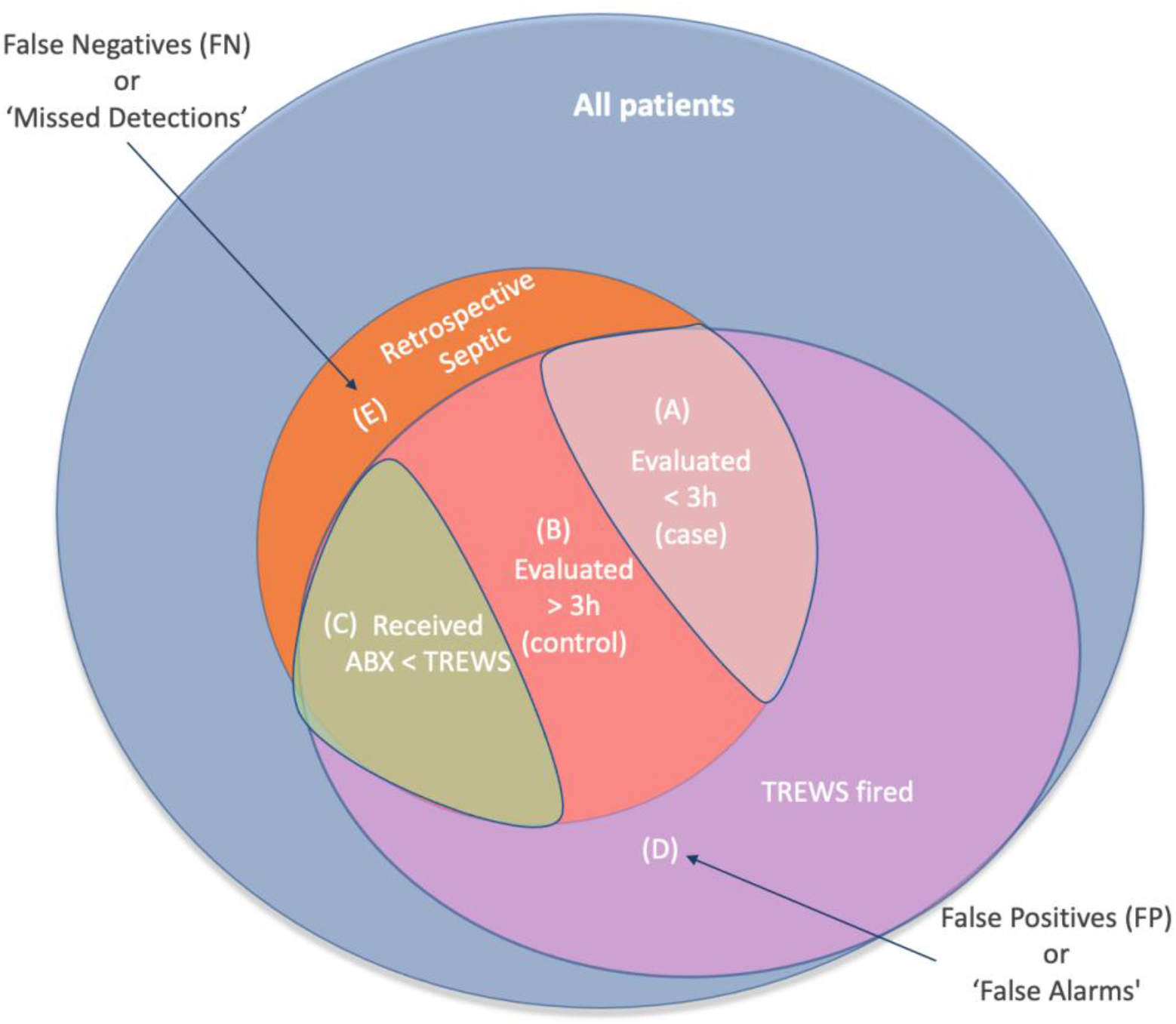
Venn diagram describing the TREWS cohort^1^, including those who were retrospectively identified as having sepsis and were identified by the ML-based alert before initiation of antibiotic therapy (shaded areas A and B). No data were reported on the false positive (D) and false negative (E) groups.

## A Causal Inference Perspective

We first attempted to construct the directed acyclic graph (DAG) representing the experimental setup of the study by Adams et al. (see Figure 2). Notably, the decision to evaluate a patient within three hours of the TREWS alert could be due to the ongoing ‘standard-of-care’ (i.e., a ‘coincident event’) triggered by the initial patient assessments and laboratory measurements. Conversely, any prolonged patient evaluation may be due to factors not within the control of the clinical team, such as increased patient volume, delayed laboratory results, case complexity or patient frailty^8^, or potential distractions due to false alarms produced by the TREWS algorithm (see Henry et al.^2^, Table 2). In such a setting, the experimental design around assignment of study and control arms of the TREWS study allowed them to take credit for early treatments and blame the clinicians for any prolonged evaluations, without properly adjusting for ‘processes of care’-related confounders (e.g., time to obtain a serum lactate result).

**Figure 2.**
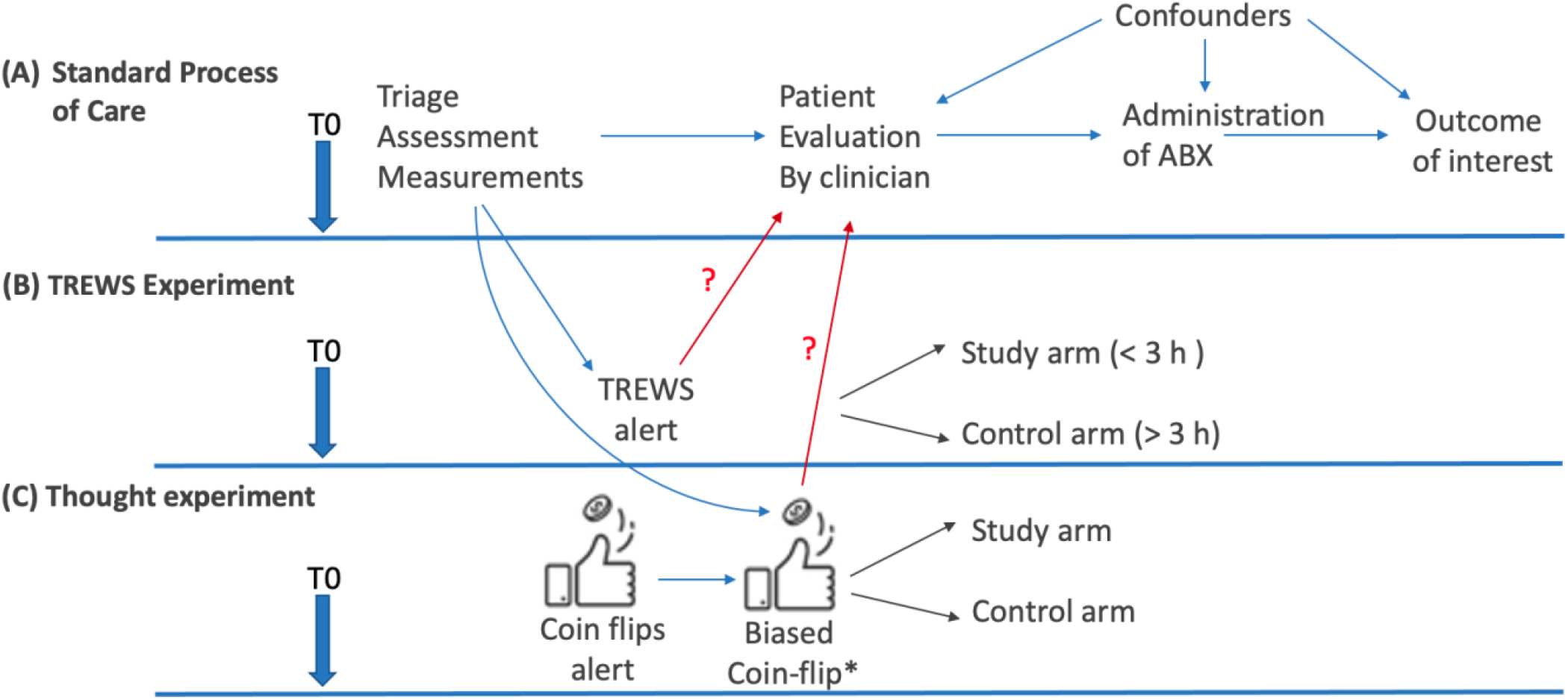
Directed acyclic graph (DAG) representations of the Standard Process of Care (A), the experimental setup of Adams et al.^1^ (B) and our proposed thought experiment (C). Note, ‘processes of care’-related factors, as well as confounders such as incoming patient volume, case complexity and frailty^8^ or distractions due to false alarms produced by the TREWS algorithm can affect patient evaluation, timing of antibiotics, and outcomes. See Appendix A for a more formal DAG representation and discussion of back-door path adjustment. ABX = antibiotics.

In the Emergency Department (ED), where the majority of sepsis cases are present on admission, we hypothesize that the actual timing of the TREWS alert is of no consequence (Hypothesis I). That is, one should be able to bucket patients into the proposed study and control arms from any random time point and arrive at similar differences in mortality. Furthermore, we hypothesize that by virtue of delaying alerts until verifiable symptoms were present^3^ (i.e., ‘looking over the shoulder’ of clinicians^5^), rather than helping to anticipate or predict cases of sepsis based on sparse or incomplete data, the TREWS alerts are increasingly correlated with the parallel and ongoing processes of care, which may alone explain the decisions to evaluate and treat patients. In such a setting, any causal claims of the potential effect of TREWS on patient outcomes may require adjustments by indicators of the processes of care (Hypothesis II); a classic *backdoor adjustment problem* in the causal inference literature^9^. As shown in Figure 2, the standard process of care triggers measurements such as labs and vitals, and the presence of certain measurements (e.g., serum lactate) are correlated with clinical suspicion of sepsis and whether the patient may receive early or late evaluation^4^. Notably, the patients in the study arm of Adams et al. were more likely to have lactate >2 mmol/L (75% versus 61%, P < 0.001)^1^. What is missing from this reporting and the subsequent risk adjustments is the number of patients who had any lactate recordings in the study vs. control arms; that is, adjustment by surrogates of processes of care and initiation of diagnostic protocols.

### The ‘Coin-flip Protocol’

To test the two hypotheses discussed above, we designed an alternative DAG in which the TREWS-based alert was replaced by a sequence of coin-flips (see Figure 2 (C)): a fair coin was flipped repeatedly, starting from the time of ED triage, and an alert was fired with the landing of the first head. This first coin flip randomly determines when an alert might be fired. Next, we used a single biased coin-flip to assign patients to either the study arm or the control arm: 60% of the patients with an available lactate >1 mmol/L at the time of the alert were assigned to the ‘study arm’, and the remaining patients (including those without a lactate measurement) to the ‘control arm’. This resulted in the study arm to include slightly higher lactate (as reported by Adams et al.^1^). Using two large patient cohorts, we assessed the impact of this study arm assignment on time-to-antibiotics and mortality. Briefly, we followed the approach of Adams et al. and included those who met Sepsis-3 criteria^7^, and received their first antibiotic after the alert and within 24 h of the alert.

Table 1 summarizes the characteristics of the patients assigned to the study and control arms according to our thought experiment (i.e., the ‘coin-flip protocol’). Notably, compared to the control arm, the study arm has slightly higher lactate, significantly lower time-to-antibiotics, lower mortality rate, lower rates of sequential organ failure assessment (SOFA) score progression at 72 h, and lower length of hospital stay. Moreover, logistic regression-based adjustment by age, gender, Charlson Comorbidity Index (CCI), serum lactate, systemic inflammatory response syndrome (SIRS), and SOFA show that the odds of mortality increased by 2.21 (p<0.001; cohort A) and 1.6 (<0.01; cohort B) for patients within the control group. In comparison the odds associated with one unit increase in SOFA is 1.30 (p<0.001; cohort A) and 1.2 (p<0.001; cohort B). However, we cannot make any claims regarding the mortality reduction benefits of our proposed ‘coin-flip protocol’, since the overall mortality for our patient cohorts remain unchanged. But, we can see that assigning patients with sepsis to different groups based on lactate availability and level results in groups with substantially different care characteristics and outcomes. As we use real-world data, these differences in care are based on information that is already available to clinicians without ANY machine learning guidance and from a randomly selected alert time. That is, the observed differences in the mortality rate are artifacts of the way patients are assigned to the study and control arms, and any alleged overall mortality reduction claims are an illusion induced by our study’s experimental setup.

### From TREWS to True Estimates

As one might expect, across the two patient cohorts analyzed in our study, the observed associations between the membership in the study arm and outcomes only goes away once adjusted for the presence of lactate (i.e., backdoor adjustment^9^). Notably, Adams et al.^1^ do not adjust for such confounders in their analysis, which is likely to significantly bias their findings. We recommend all observational studies of ML-based predictive models to include directed acyclic graph diagrams to explicitly lay out assumptions surrounding the data generation processes, and to use pertinent causal inference techniques to mitigate the problem of biased effect estimates. While thoughtful experimental design, causal inference, and rigorous reporting (including both false positive and false negative rates) can strengthen the findings of the study by Adams et al, our results demonstrate the importance of rigorously designed randomized clinical studies, a hallmark of evidence-based medicine.

## Data Availability

Access to de-identified UCSD and Grady cohort may be made available via approval from UCSD and Grady Institutional Review Board (IRB).

## Acknowledgements

Dr. Nemati has received fundings from the National Institutes of Health (R01LM013998, R01HL157985, R35GM143121), and the Gordon and Betty Moore Foundation (#GBMF9052). Dr. Holder is supported by the National Institute of General Medical Sciences of the National Institutes of Health (K23GM146092). Dr. Wardi is supported by the National Institute of General Medical Sciences of the National Institutes of Health (K23GM37182).

## Contributions

S.N., G.W., R.L.O, and A.L.H contributed to the initial study design and analysis plan. S.N. and S.P.S performed the analysis. All authors contributed to the interpretation of the findings, and contributed to the final preparation of the manuscript

## Approvals

Analysis of the de-identified data utilized in our thought experiment was conducted in accordance to the approved Institutional Review Board (IRB) protocols of UC San Diego (IRB#191940; Cohort A) and Grady Hospital (IRB#110675; Cohort B), and the requirement for informed consent was waived, as the use of de-identified retrospective data does not require patient consent under the Health Insurance Portability and Accountability Act (HIPAA) privacy regulations.

## Appendix A

Causal Inference and Backdoor Adjustment

**Figure A1.**
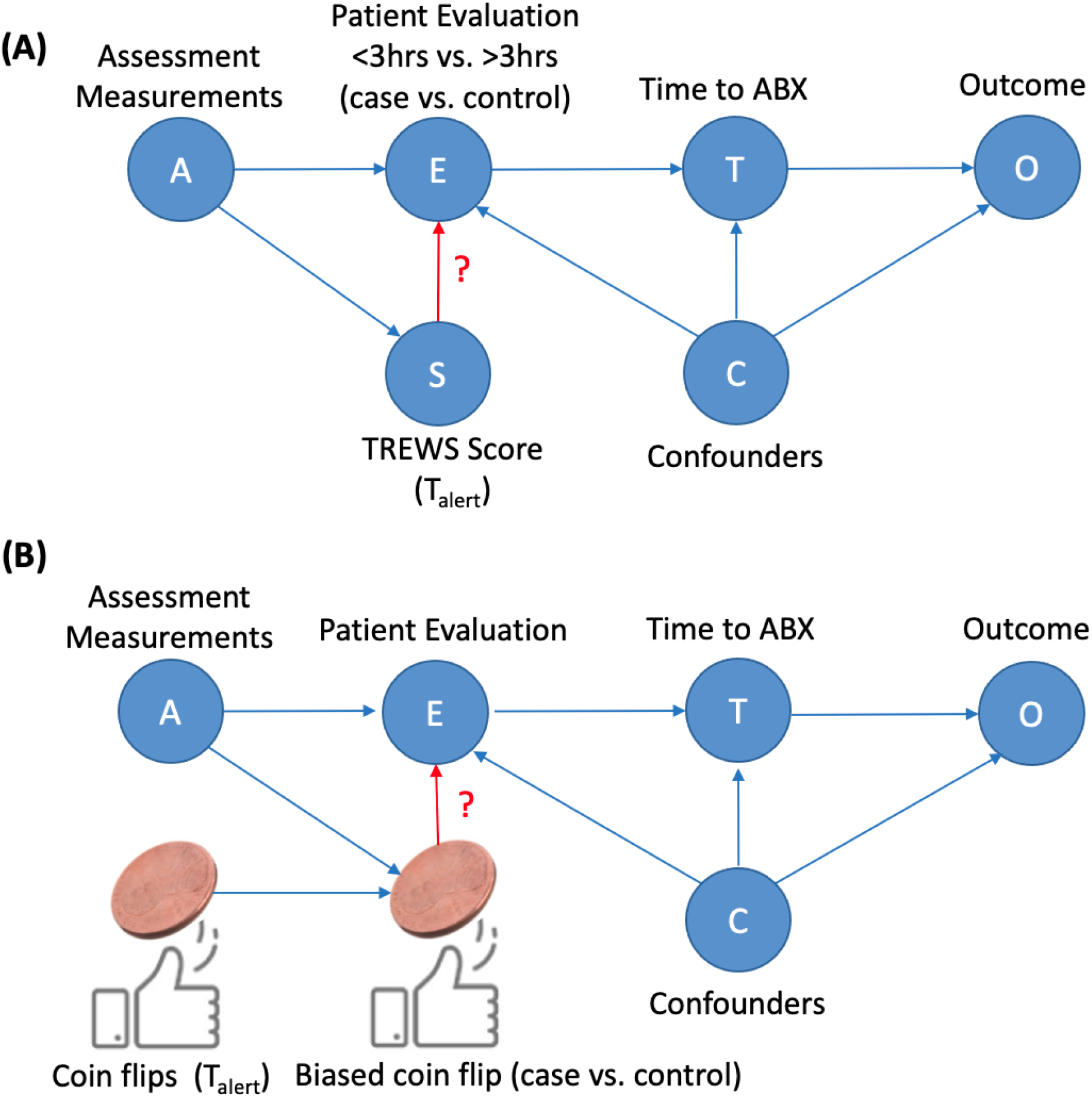
A directed acyclic graphical model representing the experimental setup of Adams et al.^1^ (top) and our proposed thought experiment (bottom).

Our thought experiment was aimed at demonstrating the confounding effect of the backdoor path induced by node A, when assessing the causal effect of node S on E, T, and O. In our thought experiment, a fair coin is flipped repeatedly, starting from the time of ED triage, and an alert is fired with the landing of the first head. Next, a second biased coin is used to assign 60% of the patients with an available lactate >1 mmol/L to the ‘study arm’, and the remaining patients (including those without a lactate measurement) to the ‘control arm’. This results in the study arm to include slightly higher lactate (as reported by Adams et al.).

The key question is whether (and under what conditions) the experimental design and the analysis method of Adams et al. can accurately estimate the effect of TREWS alerts on patient evaluation, time to administration of antibiotics and outcomes. We demonstrate that using their approach we arrive at an incorrect conclusion that a sequence of coin flips can accurately stratify patients into two groups with over S∼50% differences in mortality.

Note, ‘processes of care’-related factors (node A) (e.g., timing of a serum lactate measurement or ordering of a blood culture), as well as confounders (node C) such as patient volume, distractions caused by the TREWS false alarms, case complexity and patient frailty can affect patient evaluation, timing of antibiotics, and outcomes. However, Adams et al. do not adjust for the formers (i.e., *backdoor adjustment*), which as demonstrated in our main results (see Table 1) has the potential to substantially bias their findings. Once we control for A, we block the back-door path from S to E, producing an unbiased estimate. Similarly, in our experiment, once we adjust for the presence of serum lactate, we see that being in the study arm or the control arm has no effect on patient outcomes.

## References

1. Adams, R., Henry, K.E., Sridharan, A. et al. Prospective, multi-site study of patient outcomes after implementation of the TREWS machine learning-based early warning system for sepsis. Nat Med 28, 1455–1460 (2022). https://doi.org/10.1038/s41591-022-01894-0

2. Henry, K.E., Adams, R., Parent, C. et al. Factors driving provider adoption of the TREWS machine learning-based early warning system and its effects on sepsis treatment timing. Nat Med 28, 1447–1454 (2022). https://doi.org/10.1038/s41591-022-01895-z

3. Henry, K.E., Kornfield, R., Sridharan, A. et al. Human–machine teaming is key to AI adoption: clinicians’ experiences with a deployed machine learning system. npj Digit. Med. 5, 97 (2022). https://doi.org/10.1038/s41746-022-00597-7

4. Centers for Medicare and Medicaid Services. CMS announces update on SEP-1 validation, public reporting for Hospital Inpatient Quality Reporting. https://qualitynet.cms.gov/news/5d014bfc1543e8002ceb1d45. (2016).

5. Beaulieu-Jones, B.K., Yuan, W., Brat, G.A. et al. Machine learning for patient risk stratification: standing on, or looking over, the shoulders of clinicians?. npj Digit. Med. 4, 62 (2021). https://doi.org/10.1038/s41746-021-00426-3

6. Wang J, Strich JR, Applefeld WN, Sun J, Cui X, Natanson C, Eichacker PQ. Driving blind: instituting SEP-1 without high quality outcomes data. J Thorac Dis. 2020 Feb;12(Suppl 1):S22–S36. doi: 10.21037/jtd.2019.12.100. Erratum in: J Thorac Dis. 2021 Jun;13(6):3932-3933. PMID: 32148923; PMCID: PMC7024755.

7. Seymour, C. W. et al. Assessment of clinical criteria for sepsis for the third international consensus definitions for sepsis and septic shock (sepsis-3). JAMA 315, 762–774 (2016).

8. Lee HY, Lee J, Jung YS, Kwon WY, Oh DK, Park MH, Lim CM, Lee SM. Preexisting clinical frailty is associated with worse clinical outcomes in patients with sepsis. Critical Care Medicine. 2021 Oct 25;50(5):780–90.

9. Pearl J. Causal inference. Causality: objectives and assessment. 2010 Feb 18:39–58.

10. Sharafoddini A, Dubin JA, Maslove DM, Lee J. A new insight into missing data in intensive care unit patient profiles: observational study. JMIR medical informatics. 2019 Jan 8;7(1):e11605.

